# Mental health indicators in Sweden over a 12-month period during the COVID-19 pandemic

**DOI:** 10.1101/2021.12.10.21267338

**Authors:** Anikó Lovik, Juan González-Hijón, Anna K. Kähler, Unnur A. Valdimarsdóttir, Emma M. Frans, Patrik K. E. Magnusson, Nancy L. Pedersen, Per Hall, Kamila Czene, Patrick F. Sullivan, Fang Fang

**Affiliations:** Institute of Environmental Medicine, Karolinska Institutet, Solna, Sweden; Department of Medical Epidemiology and Biostatistics Karolinska Institutet, Solna, Sweden; Centre of Public Health Sciences, Faculty of Medicine, School of Health Sciences, University of Iceland, Reykjavik, Iceland; Department of Epidemiology, Harvard TH Chan School of Public Health, Boston, Massachusetts, USA; Department of Oncology, Södersjukhuset, Stockholm, Sweden; Departments of Genetics and Psychiatry, University of North Carolina, Chapel Hill, North Carolina,USA

**Author notes:** **Correspondence to:** Anikó Lovik, Institutet för miljömedicin, C6 Integrativ epidemiologi Fang, 171 77 Stockholm.

**Keywords:** Anxiety symptoms, COVID-19, Depressive symptoms, General population, Mental health, Omtanke2020 study, Stress, Sweden

## Abstract

**Background:** The ongoing COVID-19 pandemic has had an unprecedented impact on the lives of people globally and is expected to have profound effects on mental health. Yet, self-reported large-scale online surveys on mental health are still relatively uncommon. Here we aim to describe the mental health burden experienced in Sweden using baseline data of the Omtanke2020 Study.

**Method:** Self-reported baseline data collected over a 12-month period (June 9, 2020-June 8, 2021) from the longitudinal online survey of the Omtanke2020 Study including 27,950 adults in Sweden. Participants were volunteers or actively recruited through existing cohorts and after providing informed consent responded to monthly online questionnaires on socio-demographics, mental and physical health, COVID-19 infection, and impact. Poisson regression was fitted to assess the relative risk of high mental health burden (depression, anxiety, and COVID-19 specific PTSD).

**Result:** The overall proportion of persons with high level of symptoms was 15.6%, 9.5% and 24.5% for depression, anxiety, and COVID-19 specific PTSD, respectively. Overall, 43.4% of the participants had significant, clinically relevant symptoms for at least one mental health outcome and 7.3% had significant symptoms for all three outcomes. We also observed differences in the prevalence of these symptoms across strata of sex, age, recruitment type, COVID-19 status, region, and seasonality.

**Conclusion:** While the proportion of persons with high mental health burden remains higher than in pre-pandemic publications, our estimates are lower than previously reported levels of depression, anxiety, and PTSD during the pandemic in Sweden and elsewhere.

## 1 Introduction

Since the appearance of the Coronavirus Disease (COVID-19) in late 2019, profuse attention has been given to the mental health aspects of this disease. By late November 2021, Google Scholar had listed over 4,500,000 separate documents on COVID-19, of which more than 1,150,000 were about the mental health impact of the coronavirus pandemic. This speed of proliferation in publications on a novel topic is also unprecedented and comes with a price tag. Some of the published results feature small and/or convenience samples and present preliminary results that are difficult to generalise and hard to replicate (Nieto, Navas and Vázquez, 2020). In the first months of the pandemic, even more alarming was the retraction rate indicating ethically questionable research practices and errors that can be attributed to publication pressure as highlighted by Bramstedt (2020). For this reason, we believe it is important to report results on a larger scale while also considering the limitations of the study. In the present study, we present baseline data on the mental health of adults in Sweden collected in the Omtanke2020 Study (https://ki.se/omtanke2020) between 9 June 2020 and 8 June 2021.

## 2 Methods

### 2.1 Participants and procedure

Recruitment of the Omtanke2020 Study was open from 9 June 2020 to 8 June 2021 to all adults in Sweden (18 years or older) who have a BankID (unique electronic identification possessed by over 98% of Sweden’s adult population) and can read and understand Swedish. Participants were invited through (social) media campaigns and through personal invitation via ongoing studies at Karolinska Institutet including LifeGene (https://lifegene.se), Karolinska Mammography Project for Risk Prediction of Breast Cancer (KARMA, https://karmastudy.org) and the Swedish Twin Registry (STR, https://strdata.se). A total of 29,521 individuals participated in the study by giving informed consent, including 28,293 participants who answered at least one question. The participants were and are invited to respond to 11 monthly follow-ups after the baseline data collection. However, since all follow-ups are still open to participants, here we include only the baseline survey which was closed at the end of the recruitment period (June 2021). Figure 1 portrays the flowchart of including participants in the analysis of the present study. We included all participants who answered at least three out of the four mental health measures, namely depression, anxiety, stress, and PTSD (N = 27,950) at baseline.

**Figure 1:**
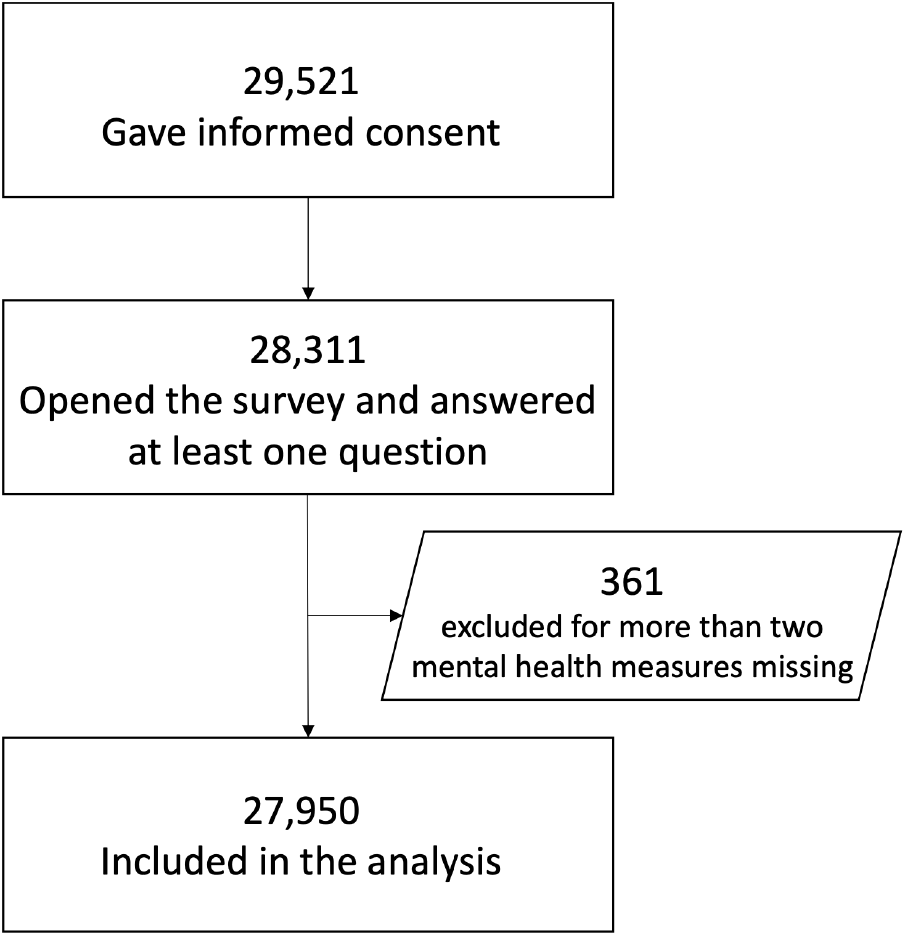
Supplementary figure 1: Flowchart of participants

### 2.2 Measures

#### 2.2.1 Demographics

We collected data on age, sex, relationship and employment status, region, and general lifestyle and health (smoking and drinking habits, body mass index, chronic diseases, and previous psychiatric disorders). We also asked participants if they joined the study through personal invitation (through a previously established cohort at Karolinska Institutet) or via our social media campaigns or other outreach activities (recruitment type).

#### 2.2.2 Mental health measures

##### Depression

The Patient Health Questionnaire (PHQ-9) is a validated screening tool for depressive symptoms and their severity, consisting of nine items measured on a four-point Likert scale (0=not at all, 1=some days, 2=more than half of the days, 3=nearly every day). Hence, the PHQ-9 total score ranges from 0 to 27. The suggested cut-off points for mild, moderate, moderately severe, and severe depression are 5, 10, 15, and 20, respectively (Kroenke, Spitzer, and Williams, 2001). The cut-off of 10 in order to create a binary outcome has been reported to have 88% specificity and 88% sensitivity. The internal consistency for the PHQ-9 based on the present study sample was *α* = 0.88.

##### Anxiety

The General Anxiety Disorder (GAD-7) scale is a validated screening tool for symptoms of generalized anxiety disorder. The scale consists of seven items measured on a four-point Likert scale as above; the total score lies between 0-21. The suggested cut-off points for mild, moderate, and severe anxiety are 5, 10, and 15, respectively (Spitzer, Kroenke, Williams, & Löwe, 2006). The cut-off of 10 in order to create a binary outcome has been reported to have 82% specificity and 89% sensitivity. The internal consistency for the PHQ-9 based on the present study sample was *α* = 0.90

##### Post-traumatic stress

We used a COVID-19-adjusted Primary Care PTSD Screen consisting of five items as a post-traumatic stress disorder (PTSD) screen for DSM V (PC-PTSD-5). The cut-off of four was chosen as having 82% specificity and 91% sensitivity (Prins, Bovin, Smolenski, Marx, Kimmerling, et al., 2016). We modified all questions to relate to COVID-19. For example, “ …have you had nightmares about the event(s) …” was replaced with “… have you had nightmares about COVID-19 …” The internal consistency for the PC-PTSD-5 based on the study sample was *α* = 0.77.

##### Stress

The four-item version of the Perceived Stress Scale (PSS-4) was used to assess stress levels on a five-point Likert-scale (Assessing the occurrence of symptoms from 1=never to 5=very often). The negatively coded (reversed) items were recoded before analysis. This scale has no established cut-off.

##### Other mental health indicators

Sleep quantity measured as hours slept per night on average during the last 14 days as well as sleep quality on a five-point Likert scale were collected (1 indicating the worst, 5 the best sleep quality). These two items were taken from the Pittsburgh Sleep Quality Index (PSQI; Buysse, Reynolds, Monk, Berman, & Kupfer, 1988). Loneliness as well as general physical and mental well-being were all measured with one item, each on a five-point Likert scale, whereas happiness was measured with one item on a ten-point Likert scale.

#### 2.2.3 COVID-19

COVID-19 status was based on self-reported, confirmed diagnosis with PCR or antibody test (data about which test was taken, when and the result were collected).

### 2.3 Statistical analyses

#### Missing data

Participants always had the response option “I cannot/do not want to answer” resulting in missing values. For demographic variables, missing values are indicated in Table 1. For validated scales, imputation based on the joint distribution was performed if the number of missing items from the scale was less than 35% (at maximum three missing items for PHQ-9, two for GAD-7, and one for PSS-4 and PC-PTSD-5). If more items were incomplete, the total score of the scale was not calculated for the participant.

**Table 1:** Socio-demographic characteristics of the partcipants

| Variable | Started<br>the survey<br>(N = 28,311) | Included<br>in analysis<br>(N = 27,950) | Tested positive<br>for COVID-19<br>(N = 2,387) | Tested negative<br>for COVID-19<br>(N = 11,887) | Not tested<br>for COVID-19<br>(N = 13,676) |
| --- | --- | --- | --- | --- | --- |
| **Sex** |  |  |  |  |  |
| Male | 5,234 (18.5%) | 5,171 (18.5%) | 491 (20.6%) | 2,242 (18.9%) | 2,438 (17.8%) |
| Female | 23,077 (81.5%) | 22,779 (81.5%) | 1,896 (79.4%) | 9,645 (81.1%) | 11,238 (82.2%) |
| **Age (years)** |  |  |  |  |  |
| Mean age (SD) | 48.6 (15.8) | 48.7 (15.8) | 44.5 (13.0) | 45.4 (14.1) | 52.3 (16.7) |
| 18-29 | 3,940 (13.9%) | 3,859 (13.8%) | 377 (15.8%) | 1,738 (14.6%) | 1,744 (12.8%) |
| 30-39 | 5,218 (18.4%) | 5,107 (18.3%) | 499 (20.9%) | 2,819 (23.7%) | 1,789 (13.1%) |
| 40-49 | 5,411 (19.1%) | 5,342 (19.1%) | 595 (24.9%) | 2,686 (22.6%) | 2,061 (15.1%) |
| 50-59 | 6,043 (21.4%) | 5,992 (21.4%) | 616 (25.8%) | 2,545 (21.4%) | 2,831 (20.7%) |
| 60-69 | 4,462 (15.8%) | 4,435 (15.9%) | 247 (10.4%) | 1,465 (12.3%) | 2,723 (19.9%) |
| 70+ | 3,237 (11.4%) | 3,215 (11.5%) | 53 (2.2%) | 634 (5.3%) | 2,528 (18.5%) |
| **Relationship status** |  |  |  |  |  |
| In a relationship | 20,500 (72.4%) | 20,234 (72.4%) | 1,800 (75.4%) | 8,668 (72.9%) | 9,766 (71.4%) |
| Single | 7,664 (27.1%) | 7,664 (27.1%) | 577 (24.2%) | 3,162 (26.6%) | 3,835 (28.0%) |
| Missing | 147 (0.5%) | 142 (0.50%) | 10 (0.4%) | 57 (0.5%) | 75 (0.5%) |
| **Employment status** |  |  |  |  |  |
| Full-time work | 14,940 (52.8%) | 14,752 (52.8%) | 1,590 (66.6%) | 7,289 (61.3%) | 5,873 (42.9%) |
| Part-time work | 2,112 (7.5%) | 2,075 (7.4%) | 188 (7.9%) | 950 (8.0%) | 937 (6.9%) |
| Not working | 2,179 (7.7%) | 2,139 (7.7%) | 172 (7.2%) | 897 (7.5%) | 1,070 (7.8%) |
| Retired | 6,895 (24.4%) | 6,836 (24.5%) | 263 (11.0%) | 1,826 (15.4%) | 4,747 (34.7%) |
| Student | 2,006 (7.1%) | 1,972 (7.1%) | 159 (6.7%) | 867 (7.3%) | 946 (6.9%) |
| Missing | 200 (0.6%) | 176 (0.6%) | 15 (0.6%) | 58 (0.5%) | 103 (0.7%) |
| **Body Mass Index** |  |  |  |  |  |
| Normal weight (<25) | 14,467 (51.1%) | 14,465 (51.8%) | 1,207 (50.6%) | 6,361 (53.5%) | 6,897 (50.4%) |
| Overweight (25-30) | 8,169 (28.9%) | 8,168 (29.2%) | 695 (29.1%) | 3,421 (28.8%) | 4,052 (29.6%) |
| Obese (> 30) | 3,707 (13.1%) | 3,707 (13.3%) | 364 (15.2%) | 1,534 (12.9%) | 1,809 (13.2%) |
| Missing | 1,968 (6.9%) | 1,610 (5.8%) | 121 (5.0%) | 571 (4.8%) | 918 (6.7%) |
| **Current smoking** |  |  |  |  |  |
| No | 22,824 (80.6%) | 22,818 (81.6%) | 1,915 (80.2%) | 9,659 (81.3%) | 11,244 (82.2%) |
| Yes | 4,662 (16.5%) | 4,662 (16.7%) | 434 (18.2%) | 2,102 (17.7%) | 2,126 (15.5%) |
| Missing | 825 (2.9%) | 470 (1.7%) | 38 (1.6%) | 126 (1.1%) | 306 (2.2%) |
| **Binge drinking** |  |  |  |  |  |
| No | 14,985 (52.9%) | 14,984 (53.6%) | 1,317 (55.2%) | 6,752 (56.8%) | 6,915 (50.6%) |
| Yes | 7,285 (25.7%) | 7,283 (26.1%) | 658 (27.6%) | 3,008 (25.3%) | 3,617 (26.4%) |
| Missing | 6,062 (21.4%) | 5,683 (20.3%) | 412 (17.3%) | 2,127 (17.9%) | 3,144 (23.0%) |
| **Somatic diseases** |  |  |  |  |  |
| None | 18,726 (66.1%) | 18,720 (67.0%) | 1,700 (71.2%) | 8,376 (70.5%) | 8,644 (63.2%) |
| One | 6,500 (23.0%) | 6,500 (23.3%) | 512 (21.4%) | 2,602 (21.9%) | 3,386 (24.8%) |
| Two | 1,731 (6.1%) | 1,731 (6.2%) | 115 (4.8%) | 617 (5.2%) | 999 (7.3%) |
| Three or more | 604 (2.1%) | 604 (2.2%) | 30 (1.3%) | 194 (1.6%) | 380 (2.8%) |
| Missing | 750 (2.6%) | 395 (1.4%) | 30 (1.3%) | 98 (0.8%) | 267 (2.0%) |
| **Previous psychiatric disorder** |  |  |  |  |  |
| No | 17,824 (63.0%) | 17,812 (63.7%) | 1,485 (62.2%) | 7,245 (60.9%) | 9,082 (66.4%) |
| Yes | 9,480 (33.4%) | 9,475 (33.9%) | 857 (35.9%) | 4,436 (37.3%) | 4,182 (30.6) |
| Missing | 1,028 (3.6%) | 663 (2.4%) | 45 (1.9%) | 98 (0.8%) | 412 (3.0%) |
| **Self-recruitment** |  |  |  |  |  |
| No | 12,125 (42.8%) | 12,122 (43.4%) | 865 (36.2%) | 4,936 (41.5%) | 6,321 (46.2%) |
| Yes | 16,207 (57.2%) | 15,928 (56.6%) | 1,522 (63.8%) | 6,951 (58.5%) | 7,355 (53.8%) |
| **Time of participation** |  |  |  |  |  |
| Jun-Aug 2020 | 7,689 (27.1%) | 7,566 (27.1%) | 494 (20.7%) | 2,083 (17.5%) | 4,990 (36.5%) |
| Sep-Nov 2020 | 8,588 (30.3%) | 8,520 (30.5%) | 413 (17.3%) | 2,876 (24.2%) | 5,231 (38.3%) |
| Dec 2020-Feb 2021 | 7,532 (26.6%) | 7,411 (26.5%) | 708 (29.7%) | 4,189 (35.2%) | 2,514 (18.4%) |
| Mar 2021-Jun 2021 | 4,523 (16.0%) | 4,453 (15.9%) | 772 (32.3%) | 2,740 (23.1%) | 941 (6.9%) |

#### Analyses

We used Spearman correlations with confidence limits to calculate correlations of the total scores of the four mental health measures and other mental health indicators (due to extreme skewness of all mental health variables). Pairwise deletion was used, with at least 27,000 observations remaining for each correlation coefficient. We used modified Poisson regression to analyse binary outcomes (as suggested in the literature, see Kroenke, Spitzer, and Williams (2001) for depression, Spitzer, Kroenke, Williams, & Löwe (2006) for anxiety, and Prins, Bovin, Smolenski, Marx, Kimmerling, et al. (2016) for PTSD), to estimate prevalences adjusted for age, sex and recruitment type unless otherwise indicated. We report the relative risk of being over cut-off (and thus experiencing significant symptoms) with 95% confidence intervals.

## 3 Results

### 3.1 Descriptives

Descriptives of the demographic characteristics among the participants included in the present study and all participants of the Omtanke2020 Study are given in Table 1. Age ranged from 18 to 94 years (mean age: 48.7 years; SD: 15.8 years) and 5,107 (18.5%) were male. All Swedish counties were represented with an over-representation of Stockholm region (12,296 persons, 44.0%). 43.4% of the participants were invited from previously established cohorts at Karolinska Institutet, mainly from KARMA (N = 5,342; 44.1%; all women), LifeGene (N = 3,592; 29.6%) and STR (N = 3,459; 28.5%) (overlaps, i.e., that a participant belongs to more than one cohort, is possible). Participation rate among those invited is 7–11%, depending on the cohort.

2,387 participants (8.5%) had been diagnosed with COVID-19 before joining the study. Table 1 also shows the descriptives of the participants depending on COVID-19 test and diagnosis status, including three groups of participants namely those who tested positive or negative for COVID-19 and those who have not taken any test before study entry. The three groups were comparable, although those who were not tested were slightly older and had a much higher percentage of retirees (35% compared to 11% and 15% for positive and negative participants, respectively). The group of COVID-19 infected participants was, in general, a bit younger and healthier (lower BMI, less smoking and binge drinking, less comorbidity) and had a high proportion of self-recruitment.

### 3.2 Mental health

Descriptives and unadjusted rank correlations of the total scores of the four mental health measures and other mental health indicators are displayed in Table 2. All these measures had at least 27,500 responses and the single-item variables had even fewer missing observations. For each measure, the whole range of possible values could be observed. PHQ-9, GAD-7 and PC-PTSD-5 were all skewed. 21,991 participants (78.7%) had minimal or mild whereas 2,550 (9.1%) had moderately severe or severe level of depressive symptoms. Anxiety was even more skewed towards low scores with 23,763 (85.1%) and 1,599 (5.7%) who had minimal and mild or severe anxiety symptoms, respectively.

**Table 2:**
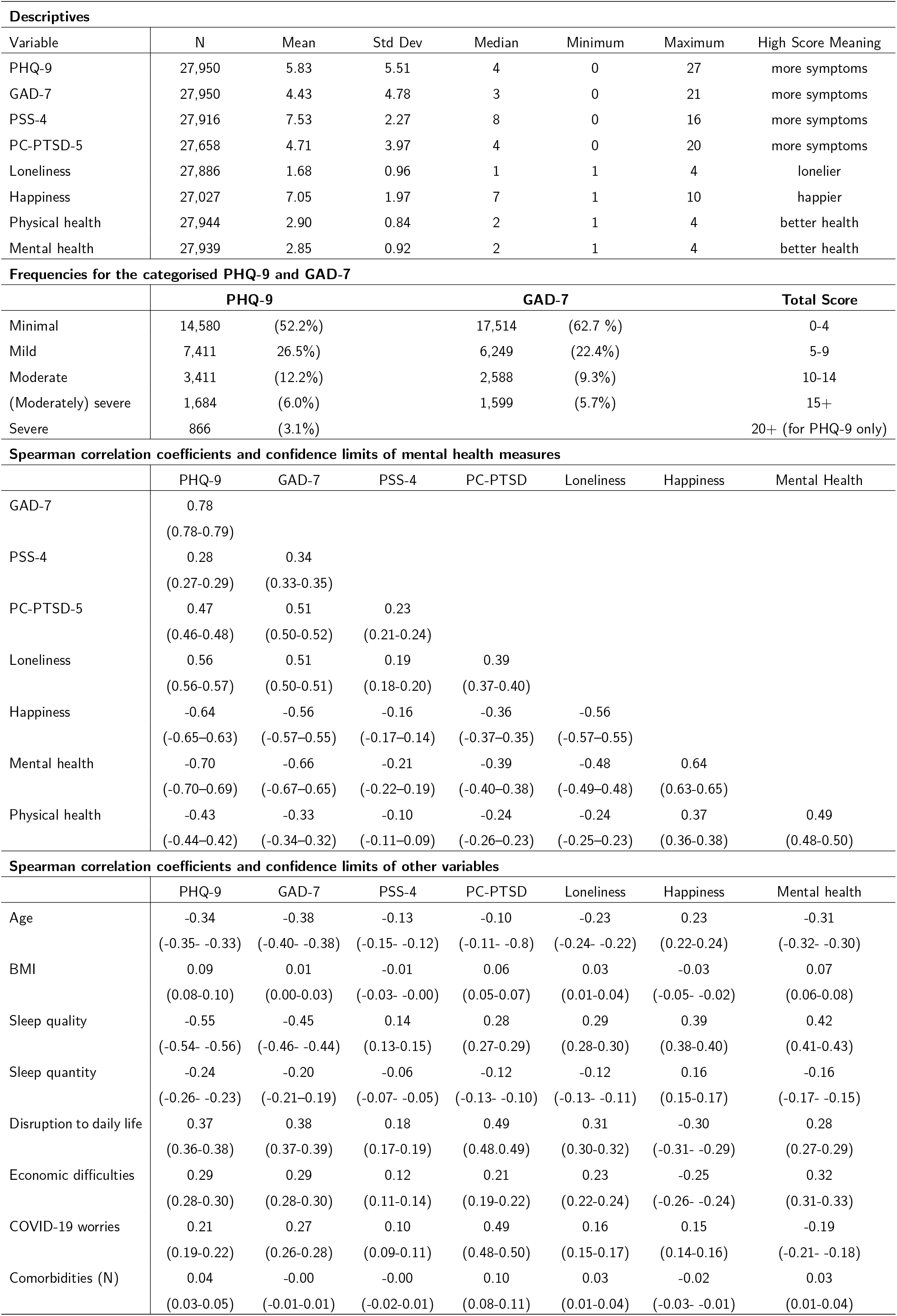
Mental health outcomes

All mental health outcomes were correlated to each other with the absolute value of the correlations ranging from 0.10 to 0.78. PSS-4 had the lowest correlations with the other measures, while PHQ-9 was highly correlated with many of the other measures, especially GAD-7 (0.78), general mental health (−0.70) and self-rated happiness (−0.64). Happiness was more strongly correlated with mental health (0.64) than physical health (0.37), whereas the correlation between mental and physical health was moderate (0.49).

Figure 2 shows the county-level mental health measures after adjustment for sex, age, and recruitment type. For PHQ-9, GAD-7 and PC-PTSD-5, the percentage of participants above cut-off level is presented, allowing comparisons not only between counties but also between outcomes. PSS-4, loneliness, and unhappiness (happiness inverted to help comparisons) with county-specific means (converted to a 0-100 scale) are also depicted. While most outcomes showed a regional difference of 2.5% (PSS-4) to 4% (unhappiness), depression symptoms seemed more varied with values as low as 14% in Blekinge to more than 19% in Gotland. Regional differences were also observed in the COVID-19-specific PTSD with a range from 20 to 27%. In general, regions with the highest prevalences in PTSD symptoms (e.g., Gävleborg, Gotland, Halland) are also the ones with high prevalences of anxiety symptoms.

**Figure 2:**
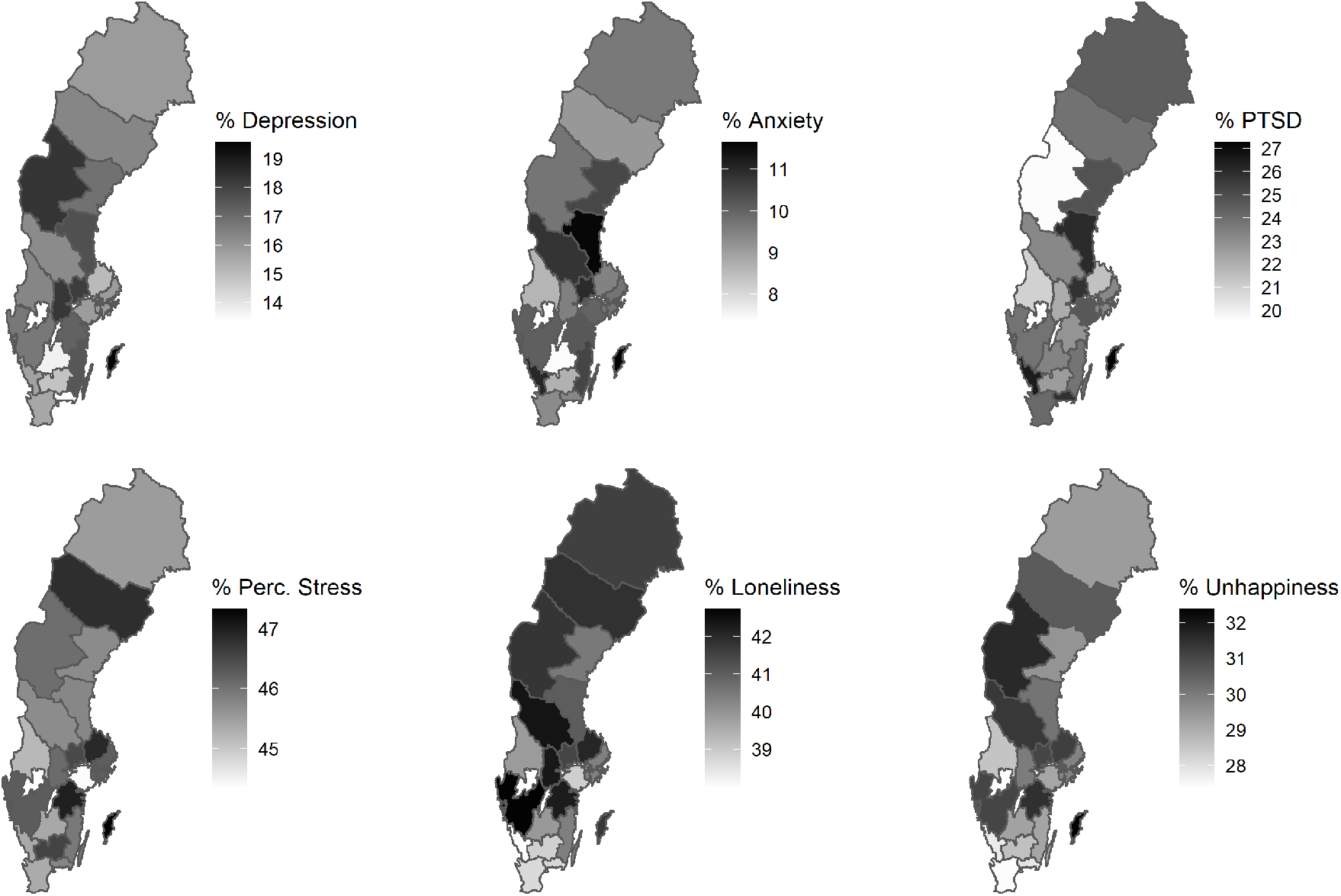
Geographical differences in mental health outcomes across Sweden

BMI, number of comorbidities and sleep quantity seemed to be less correlated to depression, anxiety, and COVID-19 specific PTSD (Spearman correlations below 0.25). Age was inversely correlated to depression and anxiety and, to a very small extent, PTSD symptoms. Perceived disruption to daily life, economic difficulties and worries about COVID-19 were all correlated to these mental health outcomes but to a varying degree (correlations range from 0.21 to 0.49). Sleep quality had a strong negative correlation with depression and anxiety but not PTSD. Perceived stress (PSS-4) was again correlated to these variables to a very low degree. For this reason and given the lack of an established cut-off for the PSS-4, the rest of the analyses focused on PHQ-9, GAD-7, and PC-PTSD-5.

Table 3 shows the relative risk of having a score above cut-off for depression, anxiety, and PTSD symptoms for the entire study sample as well as by sex, age group, recruitment type and COVID-19 status. The overall proportion of high scores was lowest for anxiety (9.5%, 4,187 participants with high level) and the highest (24.5%) for COVID-specific PTSD symptoms (8,572 participants). There were more symptoms in female participants, i.e., 3.7% and 3.3% higher prevalence of depression and anxiety, comparing female with male participants. The sex difference was greater for PTSD (12.8% higher in women). Depressive and anxiety symptoms decreased with age, although PTSD symptoms demonstrated a U-shaped form with the youngest and oldest participants scoring similarly high (27.3% and 30.0% for the youngest and oldest age groups, respectively). The biggest differences were seen between invited and self-recruited participants, where self-recruited participants had doubled relative risk of having a score above cut-off for all the three mental health measures, compared with the participants by invitation. Small differences were also observed between participants with a confirmed COVID-19 diagnosis before joining the study and others, namely that COVID-19 infected participants had a slightly higher risk of having high depressive or anxiety scores. These participants had however a lower risk of COVID-specific PTSD symptoms, compared with others (24.7% vs 22.2%).

**Table 3:** Proportion of participants with high depressive, anxiety and PTSD symptoms

|  | Depression |  | Anxiety |  | PTSD |  |
| --- | --- | --- | --- | --- | --- | --- |
| Overall proportion<sup>a</sup> | 5,961 | 15.3% (14.8%–15.9%) | 4,187 | 9.5% (9.1%–10.0%) | 8,572 | 24.5% (23.8%–25.2%) |
| Sex<sup>b</sup> |  |  |  |  |  |  |
| Female | 5,500 | 17.3% (16.8%–17.8%) | 3,598 | 11.3% (10.9%–11.8%) | 7,561 | 31.7% (31.0%–32.3%) |
| Male | 911 | 13.6% (12.8%–14.4%) | 589 | 8.0% (7.4%–8.7%) | 1,011 | 18.9% (17.9%–20.0%) |
| Age group<sup>c</sup> |  |  |  |  |  |  |
| 18–29 | 1,622 | 32.2% (30.7%–33.7%) | 1,266 | 24.0% (22.6%–25.4%) | 1,407 | 27.3% (26.1%–28.7%) |
| 30–39 | 1,457 | 22.1% (20.9%–23.7%) | 1,103 | 15.9% (15.0%–17.0%) | 1,666 | 24.6% (23.5%–25.8%) |
| 40–49 | 1,113 | 16.7% (15.7%–17.7%) | 790 | 11.3% (10.5%–12.1%) | 1,487 | 21.3% (20.3%–22.3%) |
| 50–59 | 995 | 14.6% (13.8%–15.6%) | 593 | 8.3% (7.7%–9.0%) | 1,632 | 22.1% (21.1%–23.2%) |
| 60–69 | 491 | 10.1% (9.3%–11.0%) | 284 | 5.6% (5.0%–6.3%) | 1,295 | 24.3% (23.1%–25.6%) |
| 70–94 | 283 | 8.6% (7.7%–9.6%) | 151 | 4.4% (3.8%–5.2%) | 1,085 | 30.0% (28.5%–31.7%) |
| Recruitment type<sup>d</sup> |  |  |  |  |  |  |
| Self-recruited | 4,526 | 21.1% (20.3%–22.0%) | 3,256 | 13.4% (12.7%–14.1%) | 5,773 | 31.0% (30.0%–32.0%) |
| Invited | 1,435 | 11.1% (10.6%–11.7%) | 931 | 6.8% (6.3%–7.2%) | 2,799 | 19.4% (18.6%–20.1%) |
| Confirmed COVID-19 diagnosis<sup>a</sup> |  |  |  |  |  |  |
| No | 5,336 | 15.2% (14.6%–15.8%) | 3,760 | 9.5% (9.0%–9.9%) | 12,555 | 24.7% (24.0%–25.5%) |
| Yes | 625 | 16.8% (15.7%–18.1%) | 427 | 10.4% (9.2%–11.0%) | 1,122 | 22.2% (20.8%–23.8%) |

<sup>a</sup> – Adjusted to age 50 years, sex and recruitment type; <sup>b</sup> – Adjusted to age 50 years and recruitment type;
<sup>c</sup> – Adjusted for sex and recruitment type; <sup>d</sup> – Adjusted to age 50 years, and sex

In the analysis of seasonality and potential time-effects (Figure 3), we calculated the proportion of persons above cut-off plus confidence intervals per month, after adjustment for age, sex, and recruitment type. Depressive symptoms showed the highest variation (10.5% in October 2020 vs 24.4% in June 2020 and 23.4% in June 2021) while PTSD symptoms were the most stable (21.3% in October 2020 to 29.6% in July 2020). The drop in depressive symptoms during fall 2020 coincided with the less restrictive period between the first and second waves of the COVID-19 pandemic in Sweden when gatherings up to 50 persons were allowed and the daily incidence of new COVID-19 cases and fatalities were also low. In contrast, there seems to be a worsening of mental health symptoms before Christmas (from November to December) when restrictions were in place. This is especially visible for depression where prevalence increased from 11.0% (95%CI: 9.8%-12.5%) to 17.5% (95% CI:16.7%-18.5%). The time of cohort participation (i.e., when the cohort members were invited to participate) does not seem to have an impact on the mental health estimates but affects the width of the confidence intervals due to differing sample sizes each month.

**Figure 3:**
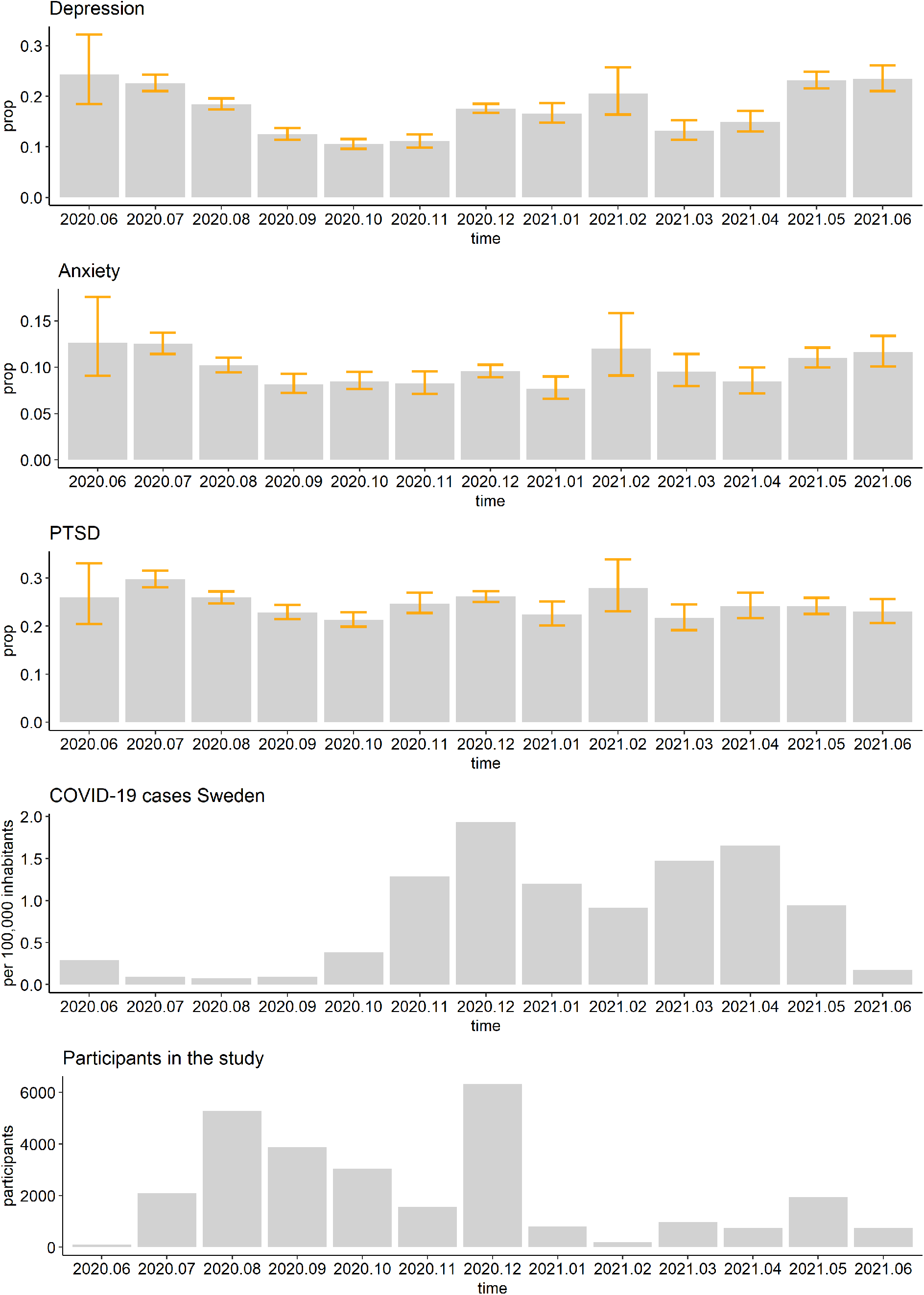
Proportion of participants above cut-off for depressive, anxiety and PTSD symptoms, COVID-19 monthly incidence per 100,000 inhabitants in Sweden, and number of new participants joining the Omtanke2020 study per month

Figure 4 shows the pattern of comorbidities between the three mental health measures. 11,725 participants (43.4%) had at least one mental health score above the cut-off. Among those with elevated values, the most common was pure PTSD symptoms (4,987 persons, 18.0%) followed by a high score for all three measures (2,036 persons; 7.4%). The least common was pure anxiety symptoms (381 persons, 1.4%) followed by anxiety plus PTSD symptoms (488 persons, 1.8%). The Venn diagrams based on age, sex and COVID-19 status all show similar proportions for the profiles with only minor differences. The most striking discrepancy was observed by recruitment type, namely that self-recruited participants had more mental health symptoms than participants by invitation.

**Figure 4:**
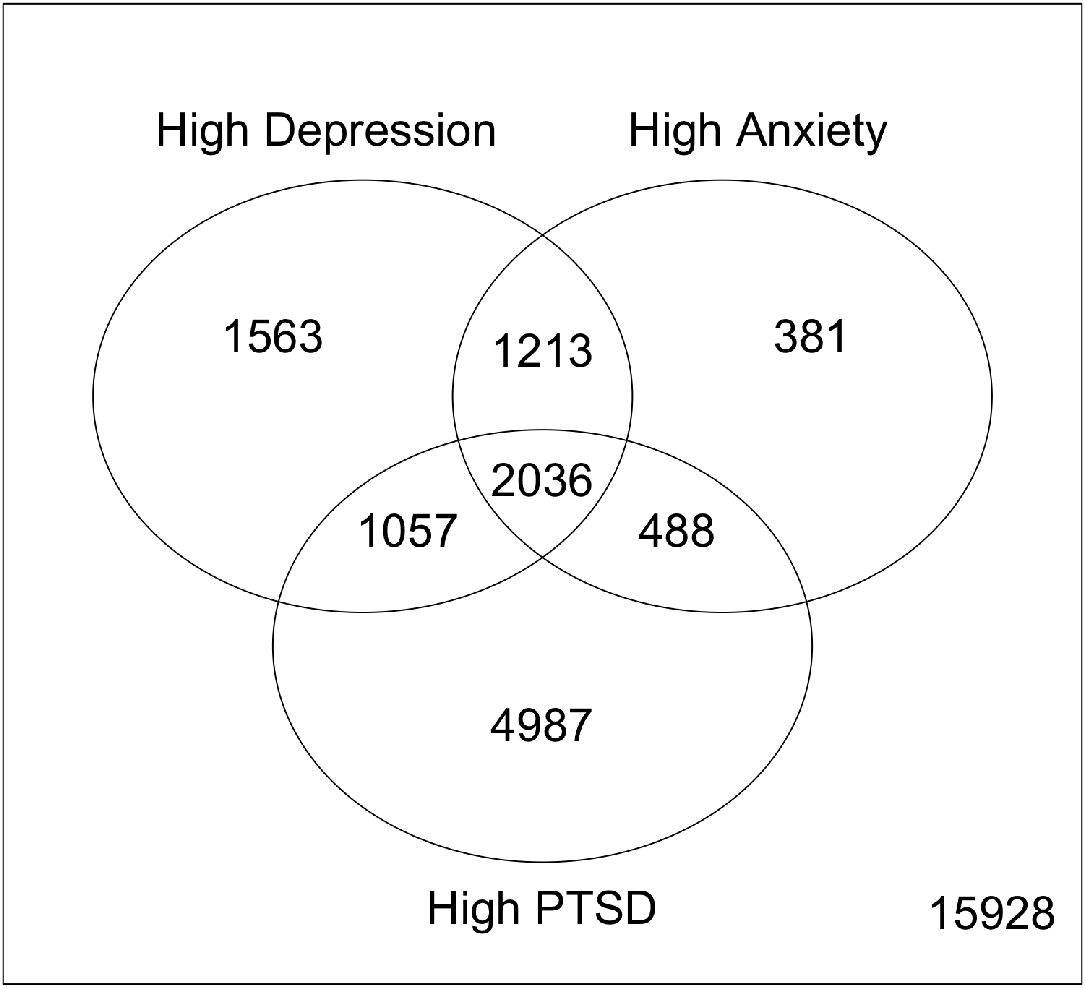
Venn diagram of the different profiles of participants with mental disorder symptoms

## 4 Discussion

In the present paper, we aimed to describe the mental health of adults in Sweden over a 12-month period during the ongoing COVID-19 pandemic. We focused on depressive, anxiety and COVID-19-specific PTSD symptoms and provided prevalence estimates overall, by sex, age group, recruitment type, COVID-19 status, region, and calendar time. Compared to what has been reported during Spring 2020 in Sweden (McCracken, Badinlou, Buhrman, & Brocki, 2020), our results suggest more moderate proportions of depressive (15.3% vs 30%) and anxiety (9.5% vs 24.2%) symptoms. This difference could have arisen both from the difference in study period (Spring 2020 vs June 2020-June 2021) and data collection method (recruitment through social media only vs recruitment through both social media and personal invitation). We demonstrated that participants recruited through social and traditional media outlets tended to have higher burden of mental health symptoms. We also found that women and younger participants had a higher prevalence of depressive, anxiety and COVID-19 specific PTSD symptoms, which is supported by a large number of studies and meta-analyses (for example, Bäuerle, Teufel, Musche, Weismüller, Kohler et al., 2020; Daly, Sutin, & Robinson, 2020; Pieh, Budumir, & Probst, 2020; Xiong, Lipsitz, Nasri, Lui, Gill, et al, 2020).

### 4.0.1 The results in context

The pre-pandemic levels of depression in Sweden were estimated as low as 8.75% (95% CI: 8.0%–9.5%, Arias-de la Torre, et al., 2021) and 10.8% (95% CI: 9.1%–12.5%, Johansson et al., 2013). Anxiety is hard to compare directly since the study by Johansson et al. (2013) used a lower cut-off (eight instead of ten) for the GAD-7 and estimated the prevalence of anxiety as 14.7% (95% CI: 12.7%-16.6%). With different methodology, Munk-Jørgensen and colleagues (2006) obtained prevalence estimates of 5.6% for anxiety among both males and females (95% CI: 3.4–7.7 for men and 3.9%–7.2% for women) but reported similar prevalence for depression as seen above (7.2% for men, 11.8% for women). Pre-pandemic PTSD-levels were estimated as 5.6% (Frans, et al., 2004). In conclusion, we have observed in general slightly higher prevalence in depression, anxiety, and PTSD in the present study, compared with pre-pandemic levels.

To put the obtained prevalences into context, we compared our results to what has been reported in other countries. Bäuerle and colleagues (Bäuerle, et al., 2020) obtained responses from over 15,000 German residents between March and May 2020 and estimated the unadjusted prevalences of depression and anxiety at 14.3% (PHQ-2, no CI reported) and 16.8%, respectively. Pre-pandemic prevalence of depression in Germany was estimated at 9.7% by Arias-de la Torre and colleagues (2021). Moreover, the unadjusted prevalence showed very similar sex differences (15.1% and 12.2% for depression and 18.3% and 10,6% for anxiety among women and men, respectively) and decreasing trend in the prevalence of both depression and anxiety by age. In Austria, based on a sample of 1,005 adults, the unadjusted prevalence was 21% for depression (PHQ-9) and 19% for anxiety (GAD-7) during the lockdown period in April 2020 (Pieh, Budimir and Probst, 2020). A large-scale meta-analysis including a sample size of 146,139 adults from worldwide data collected in Spring-Summer 2020 by Liu et al. (2021) resulted in similar estimates: 27.6% (95% CI: 24.0%-31.1%) for depression and 32.6% (95% CI: 29.1%-36.3%) for anxiety. Moreover, they also estimated a prevalence of PTSD at 16.7% (95% CI: 27.5%-36.7%). The much higher estimates in this study may be partially attributed to (1) having included studies of severely ill COVID-19 patients as well as front-line health-care workers, and (2) that many of the studies were conducted during the peak of the first wave when the mental health burden was reported to be the highest. In a longitudinal study, Daly, Sutin and Robinson (2020) found that from Spring to Summer 2020, the prevalence of mental health problems measured with the GHQ-12 decreased over time but remained significantly above pre-pandemic levels. A comparison to other Nordic countries can be found in a recent publication by Unnarsdóttir et al. (in press), which reported age- and sex-adjusted prevalence of depression as 7.6% (95% CI: 7.4%-7.9%) for Denmark, 16.6% (95% CI: 16.0%-17.2%) for Iceland, and 17.1% (95% CI: 16.1%-18.1%) and 4.2% (95% CI: 4.0%-4.3%) for two very different Norwegian cohorts.

Few studies in the current COVID-19 literature focus on the comorbidities of mental health disorders during the pandemic. A preprint from Nepal (Sigdel, Bista, Bhattarai, Pun, Giri et al, 2020) reported a prevalence of 23.2% for depression-anxiety comorbidity during April 2020. In this study, the prevalence estimates for depression and anxiety were 34.0% and 31.0%, respectively, reasonably close to the pooled estimate from Liu et al.’s meta-analysis (2021) concerning a similar calendar period. This is a much higher prevalence than the 11.6% we observed (3,249 participants, Figure 4). However, proportionally Sigdel and colleagues found less depression-anxiety comorbidity than we did, in other words in Nepal having only high depressive or high anxiety symptoms was more common than in Sweden. An Irish study on COVID-19-related PTSD reported a prevalence of 17.7% (95% CI: 15.4%-20.0%), and found very high comorbidity between PTSD symptoms and anxiety (49.5%), as well as between PTSD symptoms and depression (53.8%) (Karatzias, Shevlin, Murphy, McBride, Ben-Ezra, et al., 2020). Since we used an adjusted questionnaire to assess PTSD symptoms, direct comparison is likely not meaningful. Regardless, we identified 3,093 persons (11.1% of all participants, 25.7% of those who had at least one outcome above cut-off level) with both high depressive and high PTSD symptoms, and 2,524 persons (9.0%, 21.0%) with both high anxiety and high PTSD symptoms, also indicating high levels of comorbidity.

### 4.0.2 Strengths and limitations

The strengths of the study include a complex and inclusive recruitment strategy, allowing interested people enough time to join the study and regular (monthly) follow-ups throughout the second and third waves of the COVID-19 pandemic. We also have a much larger sample size than previous studies from Sweden (close to 28,000 compared to 1,503 in Rondung, Leiler, Meurling, & Bjártå (2021) and 1,212 in McCracken, Badinlou, Buhrman, & Brocki (2020)). Our study also has limitations. First, while the survey has been open to all, selection bias cannot be neglected. More than 80% of the sample were women which is partly because women are more likely to participate in survey research but also because the KARMA cohort (contributing over 5,500 participants) is made up entirely by women. Moreover, the younger age groups were under-represented. Further, recruitment type (through personal invitation or self-recruitment through social media and other outreach activities) was strongly related to mental health outcomes. For this reason, we corrected all analyses and figures for these factors (age, sex, and recruitment type). However, leftover influence of these factors might still have impacted the results to some extent. Regardless, the fact that over 43% of the participants scored above cut-off values of at least one of the mental health measures indicates the notable mental health burden of COVID-19. This is very close to the 45.6% reported by McCracken and colleagues, although their third outcome was insomnia (McCracken, Badinlou, Buhrman, & Brocki, 2020).

### 4.0.3 Other considerations and future perspectives

A mention of the modified PTSD scale, which now reflects the posttraumatic stress related specifically to COVID-19, is also relevant. We chose the higher cut-off of four because the item about thinking about the traumatic event (“Tried hard not to think about COVID-19”) is something 2/3 of the participants experienced at least occasionally, inflating the percentage of participants above cut-off. However, changing questions in a validated questionnaire carries a risk and further evaluation of the COVID-19 specific PC-PTSD-5 is necessary.

We also observed low correlation between PSS-4 and other mental health outcomes, including the COVID-19 specific PTSD scale. There could be several reasons for this. First, this is an ordinal measure consisting of the total score of the four items, usually compared to normative values which are not available for Sweden tour knowledge. Second, the four-item version is the short form of the original ten-item scale and short forms tend to have worse psychometric properties compared to the longer surveys (e.g., Kruyen, Emons, & Sijtsma, 2013). Third, the reliability of this four-item measure was rather low in our sample indicating that this instrument may not behave as expected. As a result, it was not included in later analyses.

We found mild to moderate correlations between economic difficulties and the studied mental health outcomes, in contrast to earlier results from Sweden by Rondung, Leiler, Meurling and Bjärtå (2021). Being worried about COVID-19 also had little effect on the mental health measures, except for the COVID-19-specific PTSD symptoms. Underlying reasons for such differences need further research. As depression and anxiety have a relatively strong link with sleep quality, future work is also needed to unravel the roles of these mental health outcomes on sleep. Further, more effort is needed to examine and validate the regional differences in mental health burden in Sweden, as well as in other countries. Understanding the underlying reasons for regional differences might be essential to understand how to mitigate the mental health burden of the pandemic and set up adequate intervention programmes.

## 5 Conclusion

In summary, we presented baseline data of our newly established Omtanke2020 Study, collecting data from nearly 28,000 adult participants. While the proportion of persons with high mental health burden remains higher than observed before the pandemic, our estimates are considerably lower than previously reported. Further, the mental health burden measures seem to follow the burden of COVID-19 pandemic to some extent.

## Data Availability

All data produced in the present study are available for a specified purpose after approval by the institution and the principal investigator of the Omtanke2020 study and with a signed data access agreement.

## Acknowledgment

This study was supported by NordForsk (project No. 105668) and Karolinska Institute (Strategic Research Area in Epidemiology and Senior Researcher Award). We acknowledge The Swedish Twin Registry for access to contact information to participating twins. The Swedish Twin Registry is managed by Karolinska Institutet and receives funding through the Swedish Research Council under the grant no 2017-00641.

## Conflict of interest

The authors declare no conflict of interest.

## Ethics Approval

The Omtanke2020 study has been registered and approved (DNR 2020-01785) by Etikprövningsmyndigheten (Swedish National Ethics Board) on 3 June 2020.

## Notes

### Competing Interest Statement

The authors have declared no competing interest.

